# Community Health Worker Outreach to Assess Confounders of Health

**DOI:** 10.1101/2024.10.31.24316456

**Authors:** Armin Takallou, Emily Mitchell, Gina M. Richardson, Brian Frank

**Affiliations:** Oregon Health and Science University, School of Medicine, Portland, OR; Oregon Health and Science University, Department of Internal Medicine, Portland, OR; University of California – San Diego, Department of Pediatrics, San Diego, CA; Oregon Health and Science University, Department of Family Medicine, Portland, OR

## Abstract

**Background and Objectives:** In July 2020, recognizing the potential conflict between COVID-19 quarantine guidelines and other medical and social needs, a university-affiliated family medicine clinic developed a workflow to support patients who test positive for COVID-19. The workflow relies on Community Health Workers (CHWs) to call patients, identify needs, and connect them to community resources, with the goal of reducing barriers to maintaining COVID-19 isolation. The objective of this study was to understand the design, implementation, and maintenance of the workflow to provide guidance for other primary care practices interested in developing similar workflows.

**Methods:** This qualitative study was conducted in a federally qualified health center. Key informant interviews were conducted with six personnel involved in the workflow. Using a semi-structured interview guide, interviewers asked participants about the design, implementation, maintenance of the workflow, and benefits and harms. Interviews were analyzed using an immersion-crystallization approach.

**Results:** Across all workflow phases, adaptability and content expertise were reported as essential for workflow success. The main barrier was the lack of coordinated COVID-19-related workflows across the system. Delivery of whole-person care was identified as the primary benefit to both patients and the healthcare system.

**Conclusions:** Interview participants felt that integrating CHWs into the workflow benefitted patients and the clinic. For other practices interested in implementing a similar workflow, themes for success include a workflow framework built on patient-centeredness, adaptability, and the unique content expertise of CHWs.

## INTRODUCTION

COVID-19 has resulted in over one million deaths in the United States since March 2020.[1] Isolating people with COVID-19 and quarantining close contacts has been a strategy to mitigate risk of viral transmission since the beginning of the pandemic. However, the ability to isolate or quarantine is influenced by health-related social needs (HRSN), such as food, childcare, medication, and safe housing.[2,3]

Federally Qualified Health Centers (FQHCs) are community-based clinics that provide primary care services in underserved communities.[4] More than two-thirds of people served by FQHCs are insured by Medicaid or uninsured, and the majority are racial or ethnic minorities, populations with disproportionate levels of unmet HRSNs.[5] Community Health Workers (CHWs) are trusted community members who share language, culture, and other lived experiences. They provide confidential, personalized services that empower individuals and their families to address HRSNs.

The Oregon Health & Science University (OHSU) Family Medicine Clinic at Richmond (FMR) is an FQHC and patient-centered primary care home serving over 16,000 people in and around Portland, OR.[6] In July 2020, FMR staff identified patients facing barriers to COVID-19 quarantine guidelines due to unmet HRSNs and developed a workflow to assist the clinic’s pre-existing “screen and intervene” process for identifying and addressing patients’ HRSNs. In the COVID-19 workflow, CHWs call COVID-19-positive patients and screen them for HRSNs. Patients with HRSNs are connected to community resources. This qualitative study aimed to explore the development, implementation, and maintenance of the workflow, including barriers and facilitators to each. While this study was specifically in the context of COVID-19, we believe that the themes discussed are widely applicable to any clinic seeking to find more information about facilitators and barriers to creating a workflow to address the HRSNs of their patients.

## METHODS

### Data Collection

Data were gathered through key informant interviews with six FMR employees directly involved with development, implementation, and maintenance of the workflow: a behavioral health supervisor (BHS), a social determinants of heath coordinator (SDC), a quality manager (QM), two CHWs (CHW1 and CHW2), and a family nurse practitioner (FNP). Participants were approached via email, and interviews were conducted via video conferencing with participants either joining from their workplace or home. No participants declined participation or withdrew from the study, and data saturation was not applicable, given that all identified key informants were interviewed. A pilot-tested semi-structured interview guide covered two domains: 1) barriers and facilitators to the development, implementation, and maintenance of the workflow and 2) perceived benefits and harms of the workflow for patients and the healthcare system. A third domain (framework) was added to encompass foundational principles for the workflow described during interviews. Interviews lasted 60 minutes. One member of the team asked questions while another took notes and asked clarifying questions to explore emerging themes. Interviews were audio-recorded, professionally transcribed, and saved on a secure network for data management and analysis. The clinic was provided an “impact fee” to compensate for employees’ time. This study was approved by OHSU IRB# 22571.

### Analysis

*Taguette* was used for qualitative data analysis. The research team analyzed the data collaboratively using an immersion-crystallization approach and Miller and Crabtree’s five-phase analysis strategy [Table 1].[7–10] First, interviews were read by all team members for initial immersion-crystallization, identification of themes, and creation of a coding template. Next, two team members re-visited and coded each individual interview. Matrices were then created to display themes and coded text across interviews for comparative analyses as suggested by Miles and Huberman.[11] Initial classifications were shared with selected informants to confirm study findings were reasonable, a qualitative research verification step known as member checking.[12] Prominent themes were identified and consolidated into outcomes by the team.

**Table 1:** Five-Phase Data Analysis Process.

|  |
| --- |
| 1. **<u>Describing:</u>** First immersion-crystallization process: data initial read (immersion) to identify overarching pattern for organizing the data (crystallization). |
| 2. **<u>Organizing:</u>** A coding template will be created to organize data and structure the subsequent analysis.□ |
| 3. **<u>Connecting:</u>** Second immersion-crystallization cycle: coded text analyzed to identify cross-cutting patterns. |
| 4. **<u>Corroborating/legitimizing:</u>** We will seek additional data to confirm/disconfirm or refute insights. |
| 5. **<u>Representing the account:</u>** We will identify ways of sharing interpretations that are meaningful for the target audiences. |

The research team consisted of students in their dual Doctor of Medicine and Master of Public Health program and a principal investigator who is a family physician at FMR. Given that the team engages deeply with both public health training and programs and clinical medicine, the interpretations of the interviews consider both social determinants of health and clinical patient experiences.

## RESULTS

> “…what we were seeing were patients of ours who were testing positive who either were expressing [a need for], or we were finding out later didn’t actually have, the basic information or concrete materials to do what we needed them to do, which is: stay home.” – BHS

During the interviews, prominent themes emerged in the domains described above [Figure 1 and Table 2]. 227 patients screened via the workflow between July 1, 2020 and December 31, 2021. Of these, 39% (n = 89) had one or more HRSN, and information on patient demographics and category of need was collected [Tables 3 and 4].

**Figure 1.**
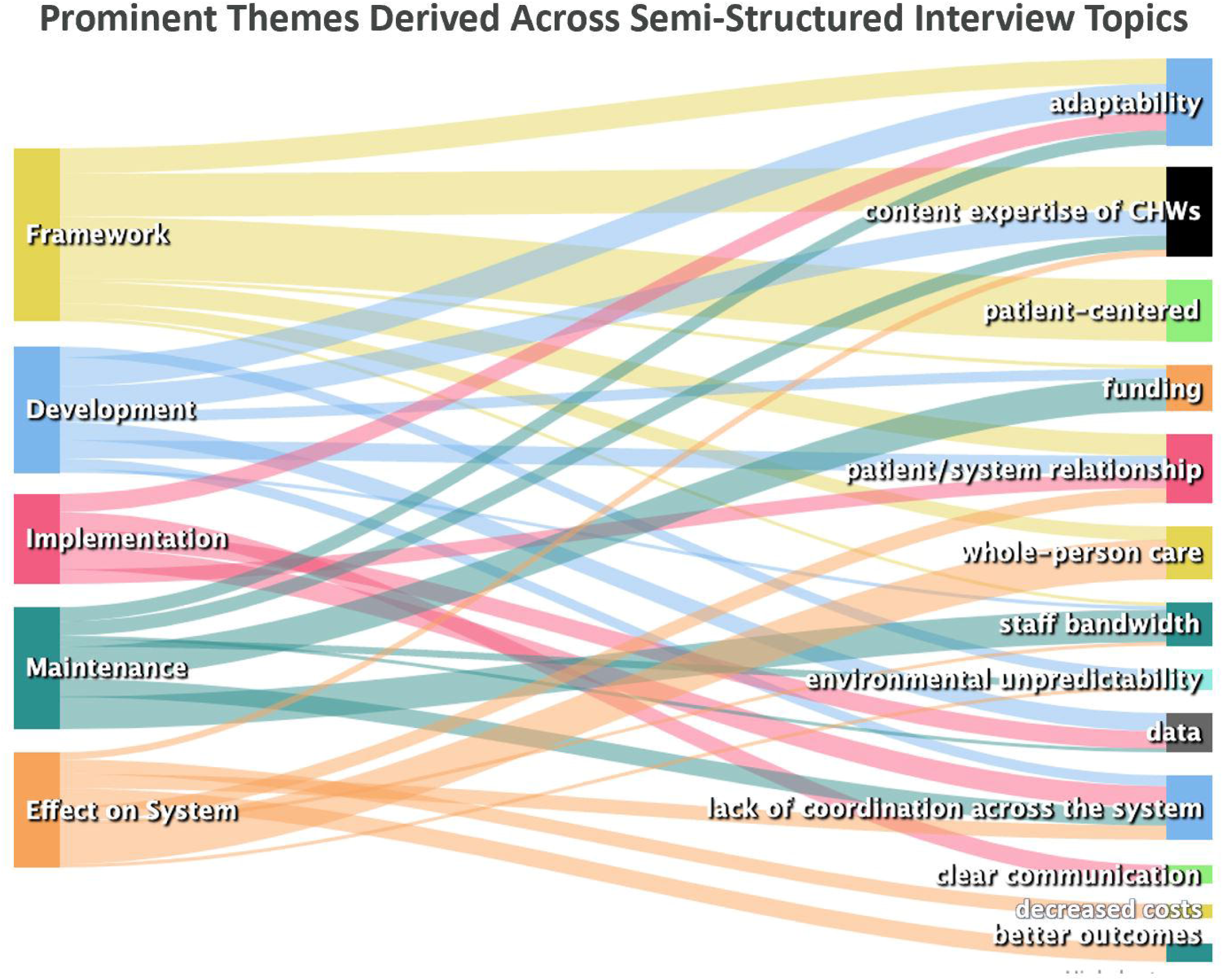
When interviewing six key personnel involved in the workflow using semi­ structured interview topics (left), prominent and overlapping themes (right) emerged across these topics.

**Table 2.** Main themes within each domain.

|  |  |  |
| --- | --- | --- |
| **Framework** | Patient-centered, whole-person care |  |
|  | **Facilitators** | **Barriers** |
| **Design** | adaptability, content expertise, funding | data, patient/system relationship, environmental unpredictability, and lack of coordination across the system |
| **Implementation** | adaptability, clear communication, and data | lack of coordination across the system and the patient/system relationship |
| **Maintenance** | adaptability, content expertise, and funding | staff bandwidth, funding, and lack of coordination across the system |
|  | **Benefits** | **Harms** |
| **Impact on the System** | whole person care, the patient/system relationship, and lack of coordination across the system | patient/system relationship, better outcomes, and decreased healthcare costs |

**Table 3:** Race and ethnicity of patients who screened positive for one or more medical or social needs during workflow outreach.

| Race |  |  | Ethnicity |  |  |
| --- | --- | --- | --- | --- | --- |
|  | N | Percent |  | N | Percent |
| AIAN | 3 | 3.4% | Non-Hispanic | 66 | 74.2% |
| Asian | 5 | 5.6% | Hispanic | 12 | 13.5% |
| Black | 16 | 18.0% | Declined | 6 | 6.7% |
| Multiracial | 1 | 1.1% | Unknown | 5 | 5.6% |
| Pacific Islander | 1 | 1.1% | Total | 89 | 100.0% |
| White | 59 | 66.3% |  |  |  |
| Declined | 3 | 3.4% |  |  |  |
| Unknown | 1 | 1.1% |  |  |  |
| Total | 89 | 100.0% |  |  |  |

**Table 4:** Medical and social needs of patients who screened positive for one or more needs during workflow outreach.

| Resource | Number of instances of need* |
| --- | --- |
| Income/Financial | 128 |
| Food | 58 |
| Housing | 47 |
| Mental health | 6 |
| Medical | 8 |
| Childcare | 8 |
| Supplies | 4 |
| Other | 5 |
| Unspecified | 21 |
| Total | 285 |

\*Some patients endorsed more than one need

### Framework

At its core, the workflow was intended to provide patient-centered care to meet patient needs in the context of their whole lives, defined by FNP as:

> “Taking care of the whole person. Not just the person sitting in front of you, the person when they go home and go back to their families. Really making sure that they can successfully stay in their community. ‘Cause that’s where people thrive.

As described by SDC,

> “[The workflow] was meant to be trauma-informed; it was meant to be person-centered, person-driven. We’ve looked at our screener and thought, ‘Okay.
>
> Which things do we need to really make sure that we ask about that would be really relevant for folks that are isolating for two weeks?’”

As with any screening, only actionable data was to be collected. SDC explained this,

> “You have to be able to support the patient in finding what they need. If you’re in a situation where you’re gonna ask someone about, ‘Hey, what do you need to be safe and isolated for two weeks?’ You have to be able to at least support them in trying to find those resources. If that is absolutely not possible, we could potentially be doing more harm to the patient-provider, patient-medical-home relationship by asking that question and not being able to answer it.”

### Barriers and Facilitators

#### Barrier: Lack of Coordination

The key barrier was a lack of coordination across the healthcare system. In addition to the FMR-specific workflow, OHSU created a statewide hotline for COVID-19-related needs. Because the hotline was also tasked with calling COVID-19-positive patients across the health system, there was frequent overlap with the FMR workflow. Receiving multiple calls was taxing for patients. BHS recalled,

> “It was sometimes irritating to be getting multiple phone calls ‘cause patients were getting calls from us, from [the hotline] …They were like, ‘I’m sick. Please stop calling me.’”

The hotline dealt with a large volume of calls which prohibited provision of individualized assistance. Though understandable, this was seen as a barrier to effective resource navigation by patients. SDC recalled,

> “they weren’t doing it in … a person-centered way and driven by the patient. I think getting a 10-page MyChart message about all the food pantries around Portland is not very helpful.”

#### Facilitator: Adaptability

The workflow team was described by members as small and nimble, able to respond quickly to the pandemic’s ever-changing landscape. This adaptability facilitated all aspects of the workflow. As mentioned by BHS,

> “I remember … some things very early on that we kept fine-tuning, which is another really nice thing about the local control of it … that we could make those changes instantly.”

SDC commented,

> “I would check in with the [CHWs] every couple of weeks and say, ‘Okay. How are things going?’ ‘What things are challenging?’ ‘What things do we need to change?’ Just being able to have that touchpoint and ability to either help adapt the program, or address whatever thing is coming up with other folks in leadership.”

The term “pivot” was used frequently. When describing patient outreach, CHW2 said,

> “You have to be able to pivot quickly based on what needs there are. We’ve developed a [template] with a little bit of scripting, but as soon as you start real life application of it, you have to be able to go off that script.”

As further evidence of the team’s adaptability, several respondents mentioned that, as cases waned and a need for vaccination outreach emerged, the team was able to refocus their efforts quickly.

#### Facilitator: Content Expertise

Another key facilitator to the workflow was the unique content expertise of CHWs, which is informed by lived experience, shared community culture, and a deep knowledge of local resources. As BHS explained,

> “[CHWs] are, in the best case, from the community, experts in community resources, experts in system navigation, and they are by design non-medicalized, so they’re not gonna be caught up in the medical piece of it. They are going to be your experts in community … They are the content experts.

Similarly, SDC commented,

> “They have not only the expertise, but the lived experience to be able to understand what things you may need to think about when you’re considering different barriers. I mean, community health work is really that lens that they bring that was necessary to understand how to approach this [social needs] assessment.”

This point was driven home by CHW1:

> <u>“[The patients]</u> were predominantly people of color who spoke other languages. Using interpreters and other things of that sort could have been challenging for people actually receiving that information. That may not have been translated correctly or people may not have even gotten a phone call.”

#### Barrier and Facilitator: The Role of Data

The electronic health record (EHR) emerged as both a barrier and a facilitator to the workflow. Early on, SDC recalled,

> “The process of how we got the information of who was COVID-positive was such a pain when it first began. I remember our previous practice manager had to go back through her emails over the last couple of weeks and just forward us emails of patient notifications of testing positive. That was a barrier. We were dependent on someone else; it was potentially disrupted by human error.”

Over time, however, the team adapted. In QM’s words,

> “We use a lot of data to inform many internal monitoring efforts. It didn’t really feel or seem like anything new, it was just like, oh, okay, we just need to work to understand the need. What are we trying to accomplish, and then, what’s the best source of data and ways of publishing it?”

An additional benefit was the ease of communicating patients’ needs to others. According to FNP,

> “Within our electronic health record, we can send messages. I would route the chart to whoever it needed to be routed to and then that person would … usually respond directly to me and say, okay, we’ll call them at this point in time. I would say they would benefit from assistance with X, Y, and Z things. They say, okay, we’ll help take care of it.”

The ease of this communication reduced barriers to tracking patient outreach for both clinical and non-clinical staff.

### Benefits of the Workflow

#### Benefits to the Healthcare System

The immediate perceived benefit of this workflow was that it improved the ability of COVID-19-positive patient to safely isolate, thereby reducing viral transmission. CHW1 observed,

> “if I’m being told I need to stay home and isolate, but I don’t have any food, I don’t have any access to go and get my medications or I desperately need to work to keep the lights on. Unless if I have someone doing this work and reaching out to me, chances are I’m gonna go back to work and I’m gonna … transmit it to other people.”

Additionally, participants felt the workflow provided a team-based approach to addressing social needs that reduced clinician burden and improved patient-centered care. SDC noted,

> “This workflow, just like a lot of other workflows that are meant to front-load barrier reduction, it takes the onus off of, for example, the provider or the nurse that is having that conversation with a COVID-positive patient to have to worry about these pieces. The clinicians can trust the fact that we got all of the data on the patients that were screened positive and did the outreach to them.”

#### Benefits to the Patient

The most important perceived benefit of the workflow to patients was its ability to address patients’ HRSN rather than simply focusing on medical care. This was seen as a contrast with traditional medical care. SDH noted,

> “We tend to be pretty prescriptive in what we want you to do as the patient. You know, ‘You are gonna take this medicine so your diabetes gets better,’ not, ‘What do you think that you need to do to make your diabetes better? What thing do you wanna focus on that might help? Is it finding food? That’s great. We can help you with that, rather than, “Here’s the pill … take the pill and go, and it’ll get better.”

FNP related a specific example:

> “I think of this couple that I saw, two Spanish speaking patients that had COVID. I was able to ask them, ‘What do you need to be successful to do this? What do you need to stay healthy?’ Being able to offer this menu of options and say, ‘Someone’s gonna reach out to you in your own language to get you the food, get you the medicines, get you all the things they need.’ They had food for maybe 24 hours. It’s so easy to be like, ‘Okay, go home. Good luck.’ That’s not what our patients need ‘cause they will get sick and they will die because of those things.”

Because it takes a patient-centered approach, another perceived benefit of the workflow was its potential to reshape patients’ relationship with the medical system. Participants shared a belief that this relationship has eroded, especially among marginalized communities. Restoring trust among patients, they reasoned, is needed to reduce health disparities. BHS summed this up, saying,

> “If our goal is to help people be healthier, live better lives, to heal our communities, then we can’t just keep saying, ‘Take this medication. Go get this test done. It doesn’t matter if you don’t have transportation. It doesn’t matter if you don’t have a refrigerator for your insulin. That’s not our business.’ We can keep doing that, and we’ll continue to have huge inequities in health, we’ll continue to have poor health outcomes and continue to lose trust … there has to be trust in these relationships if we actually wanna create any change.”

Participants saw CHWs as critical to this process. According to BHS,

> “[CHWs] already come from a restorative relationship-based lens rather than a just like, ‘Here’s a list of resources.’ Any cat can do that. They have very specific restorative and relationship-building lenses which I think made [the workflow] much easier and more successful.”

BHS added,

> “I think [this work] has to be prioritized by the system, the same way that nursing care is prioritized and physician and provider care is prioritized. I think it’s a cop-out to say, ‘Oh, well, that’s too expensive.’ “

## DISCUSSION

The COVID pandemic exploited weaknesses in partnerships between medical systems, public health authorities, and communities. In many states, CHWs were deployed to bridge these gaps because of their unique ability to form trusted community relationships. Three years later, the pandemic is transitioning to a recovery phase. Long-term effects of the virus on individuals and society are not yet known, but socioeconomic needs remain high, especially among low-income and marginalized communities. Health systems are exploring ways to screen for and address HRSN among their patients.

The use of standardized screening tools for HRSN is increasing, and many health systems have hired case managers to provide health systems and social service navigation to improve health outcomes and reduce cost drivers such as hospital readmission and emergency department utilization. Some states have also explored ways of reimbursing these services, including capitated payment arrangements or direct reimbursement of traditional health workers (a category that includes CHWs). Studies are emerging to ensure that, given the often sensitive and personal nature of the questioning, HRSN screening is patient-centered.[13]

This study examines facilitators and barriers to providing outreach, HRSN screening, and resource navigation to patients at a federally qualified health center. We found that involving CHWs throughout the design, implementation, and maintenance processes benefited both patients and the clinic. In particular, the adaptability of the workflow, and the team that deployed it, paired with the content expertise of the CHWs, was seen as instrumental to its success. Lastly, it was crucial that the workflow’s framework valued whole-person care, accomplished mainly by integrating CHWs’ perspectives and considering patients’ lived experiences.

### Limitations

This qualitative study was conducted at a single FQHC in an urban setting that had already integrated CHWs into other clinic processes, simplifying the transition to this workflow. The needs and experiences of other clinics and patient populations may vary. We also did not evaluate patients’ perspective of the service. It is possible that patients did not perceive its benefit in the same way as clinic staff. Lastly, the workflow was implemented with people attempting to isolate at home when many in-person resources were unavailable. Returning to more traditional operations may obviate much of the need for this type of workflow. Further evaluation is needed to learn whether our findings are broadly reproducible and whether fidelity to our workflow improves its adoption and maintenance.

### Conclusion

We believe that many clinics, especially those serving low-income and marginalized populations, will be interested in implementing similar workflows. Our findings can guide these clinics, helping them navigate systemic challenges and avoid potential pitfalls.

## Data Availability

All data produced in the present study are available upon reasonable request to the authors.

## Acknowledgments

The authors wish to acknowledge the contributions of the staff, faculty, and leadership of Oregon Health & Science University’s Family Medicine Clinic at Richmond.

